# Characteristics and clinical features of SARS-CoV-2 infections among ambulatory and hospitalized children and adolescents in an integrated health care system in Tennessee

**DOI:** 10.1101/2020.10.08.20208751

**Authors:** Leigh M. Howard, Kathryn Garguilo, Jessica Gillon, Adam C. Seegmiller, Jonathan E. Schmitz, Steven A. Webber, Natasha B. Halasa, Ritu Banerjee

## Abstract

**Background:** Little is known regarding the full spectrum of illness among children with SARS-CoV-2 infection across ambulatory and inpatient settings.

**Methods:** Active surveillance was performed for SARS-CoV-2 by polymerase chain reaction among asymptomatic and symptomatic individuals in a quaternary care academic hospital laboratory in Tennessee from March 12-July 17, 2020. For symptomatic patients ≤18 years of age, we performed phone follow-up and medical record review to obtain sociodemographic and clinical data on days 2, 7, and 30 after diagnosis and on day 30 for asymptomatic patients ≤18 years. Daily and 7-day average test positivity frequencies were calculated for children and adults beginning April 26, 2020.

**Results:** SARS-CoV-2 was detected in 531/10327 (5.1%) specimens from patients ≤18 years, including 46/5752 (0.8%) asymptomatic and 485/4575 (10.6%) specimens from 459 unique symptomatic children. Cough (51%), fever (42%), and headache (41%) were the most common symptoms associated with SARS-CoV-2 infection. SARS-CoV-2-related hospitalization was uncommon (18/459 children; 4%); no children with SARS-CoV-2 infection during the study period required intensive care unit admission. Symptom resolution occurred by follow-up day 2 in 192/459 (42%), by day 7 in 332/459 (72%), and by day 30 in 373/396 (94%). The number of cases and percent positivity rose in late June and July in all ages.

**Conclusions:** In an integrated healthcare network, most pediatric SARS-CoV-2 infections were mild, brief, and rarely required hospital admission, despite increasing cases as community response measures were relaxed.

**Key points:** In an integrated healthcare network in the Southeastern United States, symptomatic SARS-CoV-2 infection in children was generally mild, resolved rapidly, and rarely required hospitalization. Cases increased in children and adults as community mitigation measures became less restrictive.

## Background

Emerging reports suggest that the clinical features and outcomes associated with SARS-CoV-2 infection may have important differences across the age spectrum.[1-5] The spectrum of illness reported to be associated with SARS-CoV-2 infection in children is broad, including asymptomatic viral detection, and features of coronavirus disease 2019 (COVID-19), typically characterized by respiratory symptoms, fever, and sometimes other symptoms such as diarrhea, headache, and anosmia.[3, 6, 7] The severity of illness has been reported to range from mild, transient symptoms to illness requiring hospitalization, intensive respiratory support, and even rarely death in children. More recently, reports have emerged of a severe inflammatory syndrome occurring in the weeks after SARS-CoV-2 infection or exposure in children,[8, 9] now known as multisystem inflammatory syndrome in children (MIS-C).[10] This syndrome shares many features with toxic shock syndrome, another condition associated with multisystem organ dysfunction.[11] Some cases demonstrate clinical findings similar to Kawasaki disease, although MIS-C patients often have lymphopenia and thrombocytopenia and examination findings concerning for acute abdomen that are not typically seen in Kawasaki disease.[9, 12-14] However, current understanding of the full spectrum of manifestations of SARS-CoV-2 infection in children is very limited because most existing reports are derived from cross-sectional assessments of children with more severe illnesses, such as hospitalized children.[3, 15, 16] Less is known regarding the the symptoms and illness duration among ambulatory children in the community with SARS-CoV-2 infection. Our objective was to report the sociodemographic and clinical characteristics of all children and adolescents ≤18 years of age diagnosed with SARS-CoV-2 infection at the time of and up to 30 days after diagnosis in a large, integrated health network affiliated with an urban academic medical center in the southern United States (U.S.). We also evaluated trends in infection rates during implementation and subsequent relaxation of community mitigation measures.

## Methods

### Study design and population

Active laboratory surveillance was performed for all symptomatic and asymptomatic adults and children ≤18 years of age with a positive SARS-CoV-2 test by polymerase chain reaction (PCR) in the Vanderbilt University Medical Center (VUMC) laboratory, a quaternary care academic hospital located in Nashville, Tennessee. The primary pediatric facility in the VUMC network includes a 343-bed acute care pediatric hospital with more than 16,000 inpatient discharges, 47,000 emergency department (ED) encounters, and 360,000 ambulatory clinic encounters each year. SARS-CoV-2 testing was offered in the inpatient, ED, and ambulatory settings, which includes 9 VUMC urgent care clinics that see patients >2 years, 6 VUMC pediatric-only urgent care clinics, 2 dedicated on-campus SARS-CoV-2 testing sites, and 14 urgent care clinics located within private retail pharmacies. This network serves a sociodemographically and ethnically diverse patient population derived from across urban, suburban, and rural Tennessee, southern Kentucky, and northern Alabama. The study was reviewed and approved by the VUMC Institutional Review Board.

### SARS-CoV-2 testing

Nucleic acid amplification testing for SARS-CoV-2 was performed with a variety of methodologies with *Emergency Use Authorization* by the US Food and Drug Administration (with multiple platform-use dictated by supply limitations). These platforms included (with the percent of positive results among the total positive cohort): a modified Centers for Disease Control and Prevention (CDC) qRT-PCR assay (58.3%), the Roche 6800 (13.6%), the Hologic Panther (13.0%), the GenMark Eplex (6.3%), the Diasorin Liaison (3.9%), the Biofire FilmArray SARS-CoV-2 monoplex (3.5%), the Cepheid GeneXpert (1.2%), and the Abbott IDNOW (<1%). The CDC assay employed the N1 and N2 primer/probe sets for SARS-CoV-2 with amplification/detection on an Applied Biosystems QuantStudio 7 Flex. Specimens were nasopharyngeal or nasal swabs in viral transport media, with the exception of the IDNOW, for which dry swabs were used. Testing on the IDNOW platform was limited to asymptomatic pateints only, while the other platforms included both symptomatic and asymptomatic individuals.

### Indications and locations for testing

SARS-CoV-2 PCR testing became available in our institution on March 12, 2020. At that time, given limited testing capacity, testing criteria in children were restricted to symptomatic individuals (new onset respiratory symptoms and fever or known contact with a SARS-CoV-2 infected individual); these indications were communicated electronically to ordering providers by hospital leadership. Routine screening of all pregnant women admitted to the Labor and Delivery unit began on April 22, 2020. Routine screening of asymptomatic individuals prior to other hospital admissions, initiation of chemotherapy, stem cell or solid organ transplant, or certain surgical or medical procedures was initiated on May 4, 2020. While indications for testing expanded over time to include asymptomatic individuals admitted to hospital or undergoing outpatient procedures, testing guidance was standardized across testing sites. Daily numbers of tests performed and number of positive tests among symptomatic and asymptomatic subjects were collected from April 26, 2020 through July 17, 2020 for this assessment.

### Study follow-up

For symptomatic patients and neonates born in our hospital to SARS-CoV-2-positive women detected upon routine hospital admission, our team performed phone follow-up at days 2, 7, and 30 after the laboratory diagnosis to obtain baseline sociodemographic characteristics and ascertain data on exposures, comorbidities, clinical symptoms, and illness status. We also reviewed medical records, when available, to collect clinical and demographic information. For asymptomatic patients with a positive screening test, our team performed phone follow-up at 30 days after the positive test result.

### Statistical analysis

Descriptive analyses were conducted using STATA/SE 14.2 (StataCorp LP, College Station, TX). Daily and 7-day moving average percent positivity frequencies were calculated and stratified by asymptomatic and symptomatic status. While the majority of the analyses were done in the pediatric age group, test positivity rates were compared between children and adults during the same study period.

## Results

From March 12 to July 17, 2020, SARS-CoV-2 tests were positive in 5261/70071 (7.5%) specimens obtained at VUMC testing sites and hospitals across all age groups, including 531/10327 (5.1%) specimens in patients ≤18 years, compared to 4730/59744 (7.9%) specimens in patients >18 years.

### Asymptomatic children and adolescents

Forty-six/5752 (0.8%) specimens from asymptomatic children and adolescents screened after close contact with a SARS-CoV-2 infected individual or prior to hospitalization or medical procedures were positive for SARS-CoV-2. Among 5752 specimens tested in asymptomatic patients ≤18 years, 3766 (65.5%) specimens were tested prior to medical procedures, 1265 (22.0%) prior to hospital admission, 190 (3.3%) prior to chemotherapy, 98 for known exposure to SARS-CoV-2 infected individual, 18 (0.3%) prior to stem cell or solid organ transplant, and 415 (7.2%) for other/unknown reasons. One patient screened prior to hospital admission developed respiratory symptoms suggestive of COVID-19 and entered our ‘symptomatic’ follow-up cohort. Among 33 unique patients with initial asymptomatic SARS-CoV-2 detection during the follow-up window, none of the 10 patients with 30-day follow-up available developed symptoms suggestive of COVID-19 within 30 days.

### Neonates born to SARS-CoV-2 infected mothers

All 15 neonates born to SARS-CoV-2 infected mothers during the study period had negative PCR tests at birth and 24 hours later. One infant subsequently acquired symptomatic SARS-CoV-2 infection at 5 weeks of age, shortly after his father developed symptomatic SARS-CoV-2 infection, and entered our ‘symptomatic’ follow-up cohort. The remaining 14 neonates remained well without SARS-CoV-2 symptoms by the time of the 30-day follow-up phone call.

### Symptomatic children and adolescents

A total of 485/4575 (10.6%) specimens from 459 unique symptomatic children and adolescents were positive during the study period (**Table 1**). From these 459 children, 326 (67%) positive tests were collected in outpatient settings, 131 (29%) in the ED, and 2 (0.4%) during hospital admission. The median duration from time of symptom onset to time of SARS-CoV-2 testing was 2 days (IQR 1-3 days).

**Table 1.**
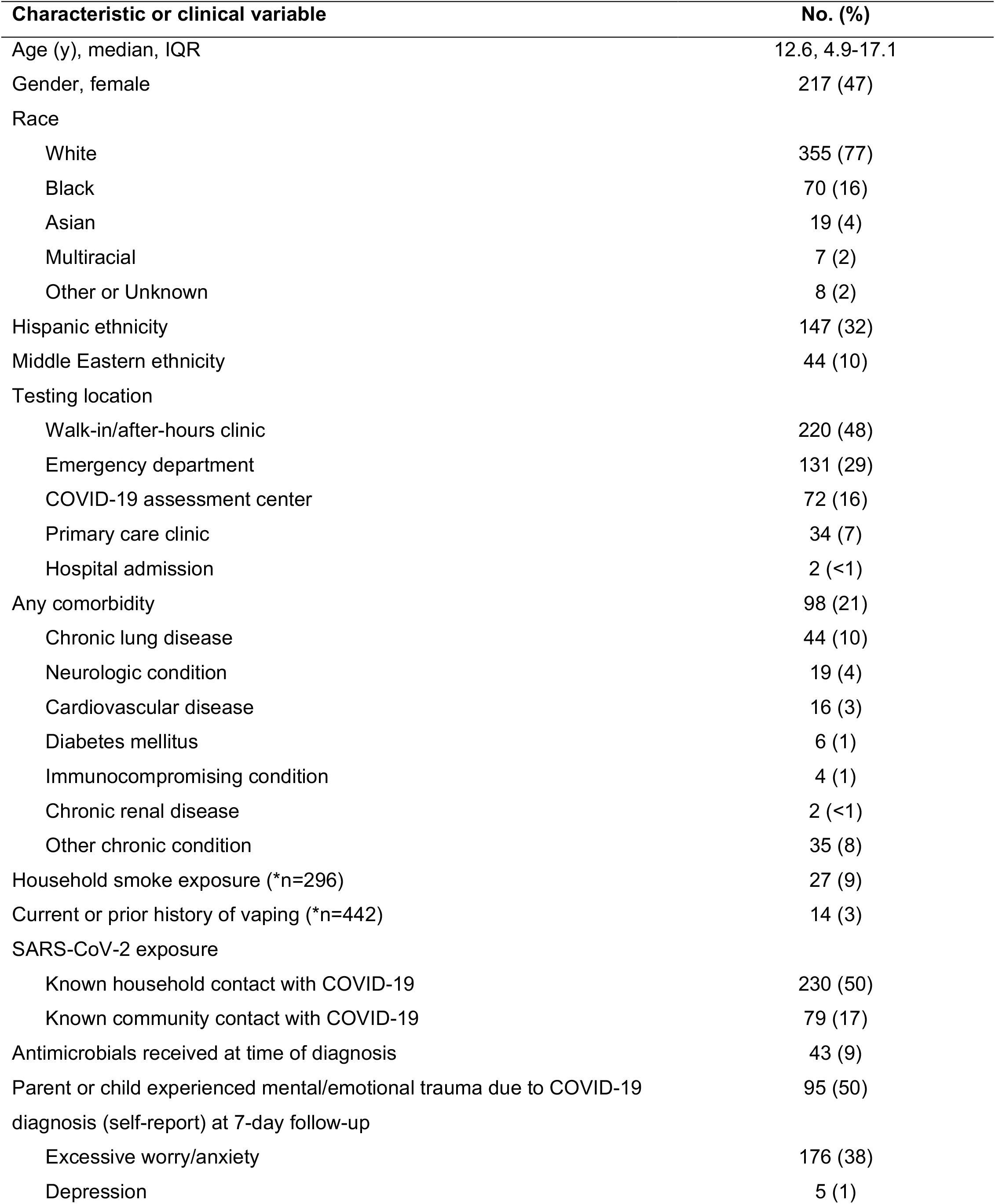
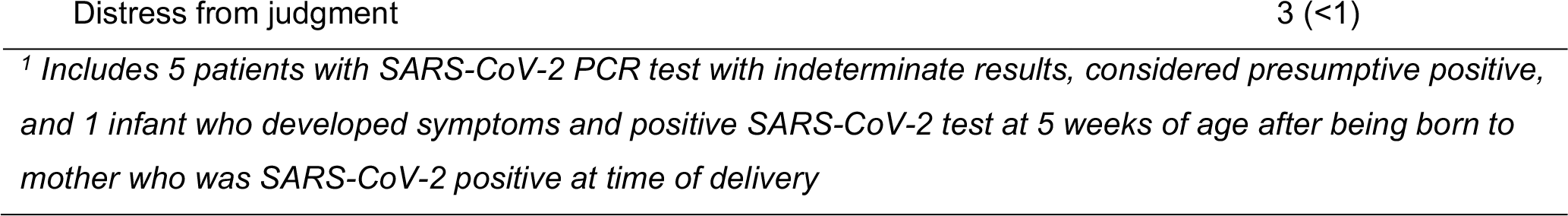
Sociodemographic and clinical features of symptomatic SARS-CoV-2 positive children and adolescents 18 years and under (n=459^1^ unless otherwise specified)

| Characteristic or clinical variable | No. (%) |
| --- | --- |
| Age (y), median, IQR | 12.6, 4.9-17.1 |
| Gender, female | 217 (47) |
| Race |  |
| White | 355 (77) |
| Black | 70 (16) |
| Asian | 19 (4) |
| Multiracial | 7 (2) |
| Other or Unknown | 8 (2) |
| Hispanic ethnicity | 147 (32) |
| Middle Eastern ethnicity | 44 (10) |
| Testing location |  |
| Walk-in/after-hours clinic | 220 (48) |
| Emergency department | 131 (29) |
| COVID-19 assessment center | 72 (16) |
| Primary care clinic | 34 (7) |
| Hospital admission | 2 (<1) |
| Any comorbidity | 98 (21) |
| Chronic lung disease | 44 (10) |
| Neurologic condition | 19 (4) |
| Cardiovascular disease | 16 (3) |
| Diabetes mellitus | 6 (1) |
| Immunocompromising condition | 4 (1) |
| Chronic renal disease | 2 (<1) |
| Other chronic condition | 35 (8) |
| Household smoke exposure (*n=296) | 27 (9) |
| Current or prior history of vaping (*n=442) | 14 (3) |
| SARS-CoV-2 exposure |  |
| Known household contact with COVID-19 | 230 (50) |
| Known community contact with COVID-19 | 79 (17) |
| Antimicrobials received at time of diagnosis | 43 (9) |
| Parent or child experienced mental/emotional trauma due to COVID-19 diagnosis (self-report) at 7-day follow-up | 95 (50) |
| Excessive worry/anxiety | 176 (38) |
| Depression | 5 (1) |

<sup>1</sup> Includes 5 patients with SARS-CoV-2 PCR test with indeterminate results, considered presumptive positive, and 1 infant who developed symptoms and positive SARS-CoV-2 test at 5 weeks of age after being born to mother who was SARS-CoV-2 positive at time of delivery

The median age of symptomatic patients was 12.6 years (IQR 4.9-17.1; **Table 1**); 60 (13%) were <1 year of age, 56 (12%) 1-4 years, 65 (14%) 5-9 years, 95 (21%) 10-14 years, and 183 (40%) 15-18 years. The majority of patients were male (53%) and white (77%); over one-third of patients were of Hispanic ethnicity or Middle Eastern ancestry. 67% had a known close contact with SARS-CoV-2 infection. Almost one-quarter (21%) of patients had at least one comorbidity.

The most common symptoms associated with SARS-CoV-2 infection were cough (51%), fever >100.4°F (42%), headache (41%), and rhinorrhea (34%; **Figure 1A**). ‘Other’ symptoms were reported at the following frequencies: nasal congestion (15%), watery or red eyes (3%), decreased oral intake, fussiness, ear pain or fullness (2%), sneezing, loss of taste, wheezing, chest pain or tightness (1%), and hypothermia, constipation, altered mental status, syncope, dizziness, possible seizure activity, and dusky discoloration of toes (<1%).

**Figure 1.**
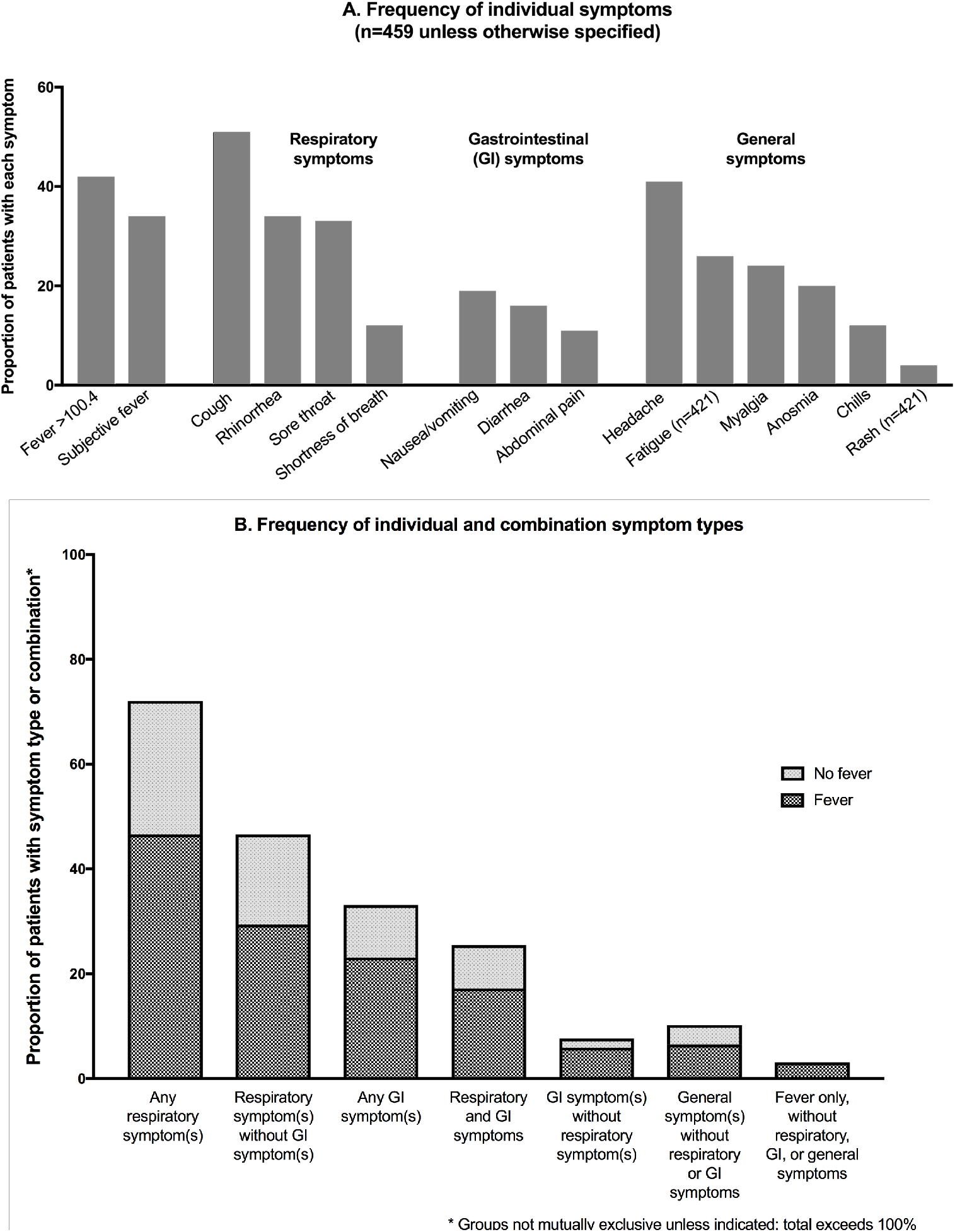
**(A)** Frequency of individual symptoms according to symptom type (fever, respiratory symptoms, gastrointestinal symptoms, general symptoms); **(B)** Frequency of occurrence of symptom types alone and in combination.

We classified symptoms as respiratory (cough, rhinorrhea, sore throat, shortness of breath), gastrointestinal (GI; nausea/vomiting, diarrhea, abdominal pain), or general (headache, myalgia, anosmia, fatigue, chills, rash; **Figure 1A**). In **Figure 1B**, the proportion of patients who exhibited respiratory, GI, and/or general symptoms, with or without fever are displayed. The majority of patients (331/459; 72%) exhibited at least one respiratory symptom, most commonly (214/331 [65%]) without GI symptom(s) present. Only 35/459 (8%) patients presented with GI symptoms in the absence of respiratory symptoms. General symptoms, without respiratory or GI symptoms, occurred in 10% of patients. Fourteen (3%) patients presented with fever without respiratory, GI, or general symptoms.

Twenty-one of 459 (4.6%) patients were hospitalized shortly after diagnosis of SARS-CoV-2 infection; 3 patients’ hospitalizations were considered unrelated to SARS-CoV-2 (scheduled chemotherapy, minor trauma, suicidal ideation) and 18 (3.9%) patients’ hospitalizations were considered possibly related to SARS-CoV-2 infection. Among these 18 patients, 13/18 (72%) were white, 5/18 (28%) were black, and 7/18 (39%) were of Hispanic ethnicity. One patient required admission twice, resulting in 19 possibly SARS-CoV-2-related hospitalizations. Median length of stay among these 19 hospitalizations was 1.5 days (range 1 to 12 days); 11/19 admissions lasted 48 hours or less. Details of these 19 hospitalizations are listed in **Table 2**. No patient required intensive care unit admission, mechanical ventilation, or extracorporeal membrane oxygenation during the study period. One patient with multiple comorbidities, not included among the 18 patients hospitalized, died upon presentation to the emergency department; it is unclear if the cause of death was related to SARS-CoV-2 infection. We observed 1 case of MIS-C during the study period.

**Table 2.** Hospitalizations possibly related to SARS-CoV-2 infection among 459 symptomatic children and adolescents up to 18 years of age

| Age<sup>+</sup> | Underlying comorbidities | Indication for hospitalization | LOS<sup>+</sup> (d) |
| --- | --- | --- | --- |
| 2 w | None | Febrile neonate | <2 |
| 7 w | None | Febrile neonate | 2 |
| 5 w | None | Brief resolved unexplained event | <2 |
| 4 m | None | Brief resolved unexplained event | <2 |
| 6 m | Complex congenital heart disease | Apneic episodes | <2 |
| 1 y | Complex neurologic condition,<br>complex congenital heart disease | (1) Respiratory distress, hypoxemia<br>(2) Cyanotic episodes | (1) <2<br>(2) <2 |
| 3 y | None | Diffuse skin desquamation, suspected<br>Staphylococcal scalded skin syndrome | 4 |
| 4 y | Complex neurologic condition | Respiratory distress, hypoxemia | 5 |
| 10 y | Hemoglobin SS disease | Acute chest syndrome | 7 |
| 12 y | Factor V Leiden | Hepatitis, presumed viral, no alternative<br>etiology identified. Resolved at 2 week<br>follow-up. | <2 |
| 12 y | Complex neurologic condition | Respiratory distress, increased seizure<br>frequency | <2 |
| 12 y | None | Dyspnea, orthostatic tachycardia | <2 |
| 14 y | B-cell acute lymphoblastic<br>leukemia | Fever and neutropenia | 12 |
| 14 y | Hemoglobin SS disease | Pain crisis | 2 |
| 15 y | Complex neurologic condition | Respiratory distress, hypoxemia | 11 |
| 17 y | Complex neurologic condition | Feeding intolerance, dehydration | 6 |
| 18 y | Hemoglobin SS disease | Pain crisis | 8 |
| 18 y | None | Hepatic abscess (culture from drainage<br>procedure grew *Streptococcus*<br>*constellatus*) | 6 |

<sup>+</sup> w=weeks, m=months, y=years, LOS=length of stay, d=days

Complete symptom resolution occurred by day 2 in 192/447 (43%) and by day 7 after diagnosis in 335/429 (78%) patients who could be reached for follow-up. Among 94 patients with ongoing symptoms at day 7, the median age was 14.4 years (IQR 6.4-17.4), and the most commonly reported ongoing symptoms were cough (34 patients), ‘other’ symptoms (33 patients), anosmia (23 patients), fatigue (18 patients), and headache (12 patients). At day 7, 177/429 (41%) patients reported experiencing excessive worry or anxiety pertaining to their COVID-19 diagnosis. Symptom resolution occurred by follow-up day 30 in 373/396 (94%) patients who could be reached for follow-up. Among 23 patients with ongoing symptoms at day 30, the median age was 12.8 years (IQR 3.7-17.4), and the most commonly reported ongoing symptoms were cough, fever, fatigue (4 patients), anosmia (3 patients), chest pain, headache, abdominal pain, ageusia, and nasal congestion (2 patients). Thirty-nine reported ongoing anxiety related to their COVID-19 diagnosis at day 30. No child was diagnosed with recurrent SARS-CoV-2 infection during the study period.

### Prevalence and percent positivity in pediatric and adult samples

Overall, from April 26 to July 17, 2020, a median of 536 specimens (interquartile range [IQR] 415-742) were tested for SARS-CoV-2 per day at our institution across all ages, which increased over the study period. The number of positive tests per day and the 7-day average percent positivity for symptomatic and asymptomatic children and adults are shown in **Figure 2**. After early variability, corresponding with periods of community mitigation measures, the number of positive tests and percent positivity began to rise in mid-June for both children and adults, corresponding to liberalization of community mitigation restrictions. Case numbers and percent positivity remained high at the end of the observation period.

**Figure 2.**
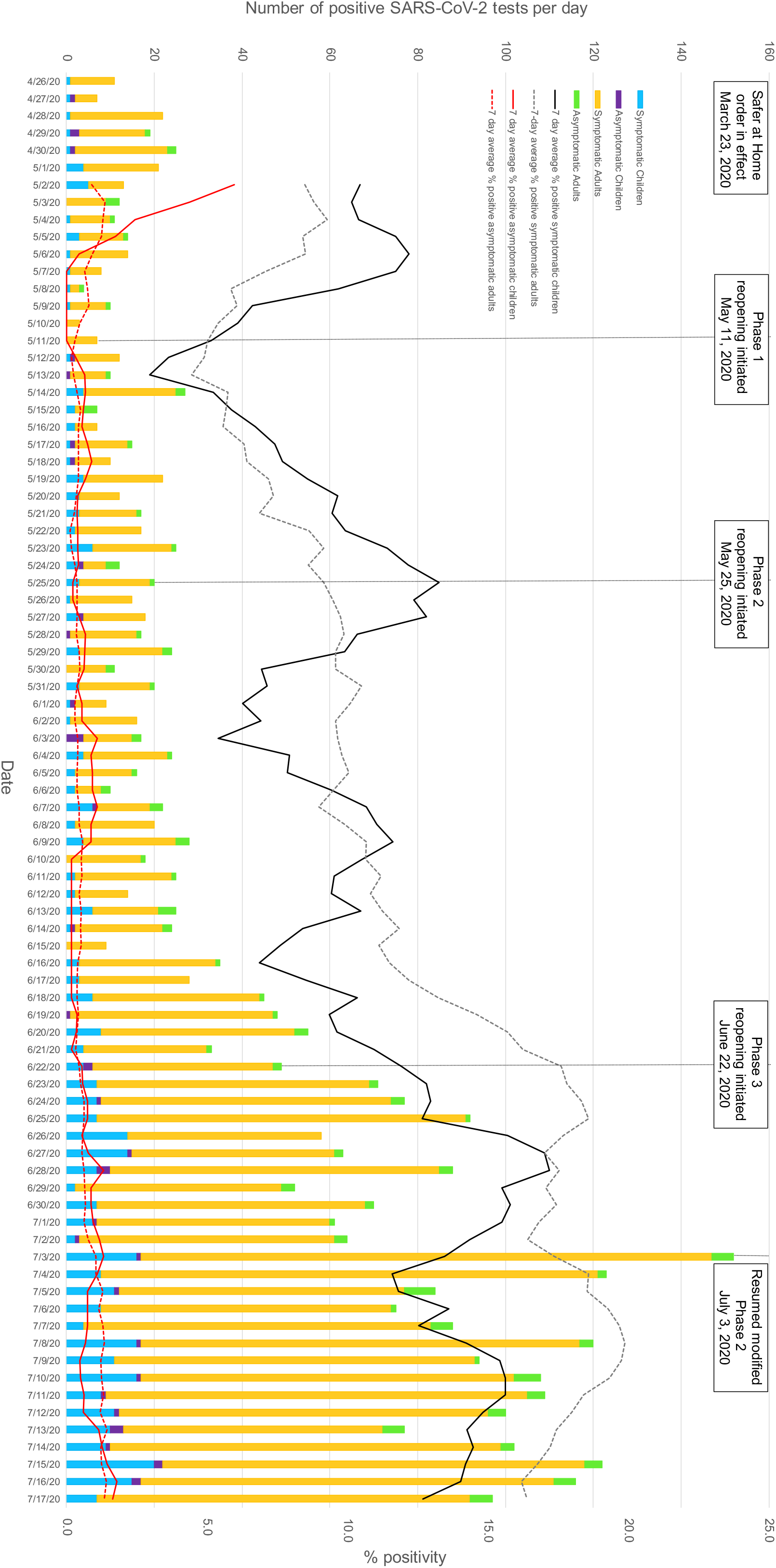
Number of samples tested for SARS-CoV-2 from April 26, 2020 to July 17, 2020 among symptomatic and asymptomatic pediatric (≤18 years) and adult (>18 years) patients (left y-axis); 7-day average percent of samples tested positive for SARS-CoV-2 (right y-axis).

## Discussion

In this cohort of children tested for SARS-CoV-2 at a single academic institution in the southeastern U.S. from March 12-July 17, 2020, the frequency of SARS-CoV-2 infection was substantially higher among symptomatic (10.6%) than asymptomatic (0.8%) children. Symptomatic children and adolescents in our cohort most commonly exhibited febrile respiratory syndromes without GI symptoms, which were generally brief and mild, with only a small minority of 459 symptomatic patients (4%) requiring hospital admission, and no patients requiring intensive care or mechanical ventilation during the study period. Despite high levels of community transmission of SARS-CoV-2 in our region, the overall prevalence of SARS-CoV-2 infections in our cohort was slightly lower in children than adults.

These findings are consistent with prior reports that children with SARS-CoV-2 infection often exhibit mild clinical presentations, and generally experience less severe clinical outcomes than adults.[1, 3, 6, 7, 16] In one of the earliest published cohorts of pediatric COVID-19 cases from the U.S., consisting of cases voluntarily reported to the Centers for Disease Control and Prevention, an estimated range of 5.7-20% of pediatric COVID-19 cases required hospitalization, while 29% of adults with SARS-CoV-2 infection required hospitalization in a separate report from an integrated health care system in California.[1] In an assessment of all hospitalized patients with SARS-CoV-2 infection in a hospital system in New York, <1% of hospitalizations occurred in children, and none died, compared to deaths occurring in 21% of hospitalized adults.[16] Recently, an overall rate of 8 hospitalizations per 100,000 children <18 years of age was reported in a population-based surveillance system comprised of data from 14 states, with approximately one-third of hospitalized children requiring ICU admission, with substantially higher hospitalization rates among Hispanic and black children than among white children.[17] Similarly, we observed that Hispanic children comprised roughly one-third of symptomatic patients and nearly 40% of hospitalized patients in our cohort. In another recent report from a large, integrated pediatric healthcare network in Pennsylvania, despite a relatively low frequency of critical illness, a higher proportion of patients required hospitalization for SARS-CoV-2-related illness (approximately 12%), respiratory support (5.7%), and mechanical ventilation (2.8%) than in our cohort. The reasons that severe illness was less common in our cohort compared to these prior studies are unclear, as baseline clinical and demographic characteristics of the patients were similar between the studies.

The clinical features of illness observed in our cohort were similar to other pediatric studies from the U.S., which reported that fever, cough, or shortness of breath occurred in 73%[3] and 75%[7] of pediatric patients, compared to 81% in our study. Another recent study of adults and children found that 96% of 164 symptomatic COVID-19 patients experienced fever, cough, or shortness of breath, and 45% exhibited all three symptoms.[18] However, this study included only 9 pediatric subjects (<18 years). We also found a slightly higher frequency of general symptoms such as headache (41%), fatigue (26%), and anosmia (20%) than in prior pediatric reports.[3, 7] Our study is unique in that we followed duration of illness and observed that the majority of children had symptom resolution within 1 week of diagnosis.

Our finding that SARS-CoV-2 was less frequently identified in children than adults has been consistently demonstrated in other studies from the U.S. and other countries.[4-6, 19] In an early report from the U.S., pediatric cases comprised <2% of all voluntarily reported COVID-19 cases prior to early April 2020.[3] Whether this represents a truly lower prevalence of infection in children, or differential testing patterns between children and adults, potentially related to milder disease phenotypes in children, is not well-established, although studies have suggested that risk of developing clinical symptoms following SARS-CoV-2 infection increases with age.[20] The implications of these findings on public health measures to mitigate transmission are still being unraveled. For other respiratory viruses, such as influenza, children are considered key drivers of transmission. However, early data suggest that children may not be the primary drivers of SARS-CoV-2 transmission within households and communities.[21] Thus, the impact of primary interventions to mitigate SARS-CoV-2 transmission aimed at children might have a relatively minor impact, particularly if SARS-CoV-2 transmissibility is low in the setting of only mild or subclinical infection.

In our community, a “Safer at Home” order went into effect on March 23, 2020, directing all residents of the county to stay home unless engaged in certain essential activities, with restrictions in place pertaining to operation and capacity of certain local businesses, including retail shops, non-essential service establishments, bars, and restaurants. A phased reopening process was initiated on May 11, 2020, involving clear guidelines for liberalizing restrictions, with built-in metrics for reversion to earlier phases as necessary. The city proceeded from Phase 1 to Phase 2 on May 25, to Phase 3 on June 22, and reverted to modified Phase 2 on July 3, 2020.[22] The increases in symptomatic cases in late June in both children and adults suggests that liberalization of these community mitigation measures may have had some impact on infection patterns.

Our study has a number of important strengths. By identifying all children with SARS-CoV-2 detections in the context of an integrated health network as the outbreak emerged, our study enabled a comprehensive assessment of the full spectrum of pediatric infection in the community, from asymptomatic infection to more severe illness. By prospectively following these subjects, we were able to assess not only the symptoms present at diagnosis, but also to follow the duration of symptoms, and to follow asymptomatic subjects for the development of symptoms after the initial detection.

Our study is also subject to some limitations. Our cohort is limited to patients who visited one of our network’s testing sites for assessment. Due to limited testing capacity, especially earlier in the study period, testing was recommended only for individuals with new-onset respiratory symptoms, fever, or a known SARS-CoV-2-positive contact; thus, providers were discouraged from sending SARS-CoV-2 testing on patients with other symptoms, such as gastrointestinal or general symptoms alone, which introduced sampling bias. Real-time monitoring of testing indications was impractical and testing practices may have varied by provider.

In summary, symptomatic SARS-CoV-2 infections among individuals ≤18 years were generally associated with respiratory symptoms, mild illness that usually resolved within a week, and rarely required hospitalization. Cases increased in our community in both children and adults as local businesses reopened, highlighting the importance of community mitigation strategies in reducing SARS-CoV-2 transmission.

## Data Availability

The data referred to in the manuscript are not currently publicly available as external datasets.

## Acknowledgments

We are grateful for the hard work and dedication of the Vanderbilt University Medical Center clinical laboratory staff and Dr. Swathi Eyyunni.

## References

1. Myers LC, Parodi SM, Escobar GJ, Liu VX. Characteristics of Hospitalized Adults With COVID-19 in an Integrated Health Care System in California. JAMA 2020.

2. Guan WJ, Ni ZY, Hu Y, et al. Clinical Characteristics of Coronavirus Disease 2019 in China. N Engl J Med 2020.

3. Team CC-R. Coronavirus Disease 2019 in Children - United States, February 12-April 2, 2020. MMWR Morb Mortal Wkly Rep 2020; 69(14): 422–6.

4. Covid-19 National Emergency Response Center E, Case Management Team KCfDC, Prevention. Coronavirus Disease-19: The First 7,755 Cases in the Republic of Korea. Osong Public Health Res Perspect 2020; 11(2): 85–90.

5. Sun K, Chen J, Viboud C. Early epidemiological analysis of the coronavirus disease 2019 outbreak based on crowdsourced data: a population-level observational study. Lancet Digit Health 2020; 2(4): e201–e8.

6. Dong Y, Mo X, Hu Y, et al. Epidemiology of COVID-19 Among Children in China. Pediatrics 2020; 145(6).

7. Otto WR, Geoghegan S, Posch LC, et al. The Epidemiology of SARS-CoV-2 in a Pediatric Healthcare Network in the United States. J Pediatric Infect Dis Soc 2020.

8. Riphagen S, Gomez X, Gonzalez-Martinez C, Wilkinson N, Theocharis P. Hyperinflammatory shock in children during COVID-19 pandemic. Lancet 2020.

9. Verdoni L, Mazza A, Gervasoni A, et al. An outbreak of severe Kawasaki-like disease at the Italian epicentre of the SARS-CoV-2 epidemic: an observational cohort study. Lancet 2020.

10. Prevention CfDCa. Multisystem Inflammatory Syndrome in Children (MIS-C) Associated with Coronavirus Disease 2019 (COVID-19). Available at: https://emergency.cdc.gov/han/2020/han00432.asp. Accessed May 19, 2020.

11. Lappin E, Ferguson AJ. Gram-positive toxic shock syndromes. Lancet Infect Dis 2009; 9(5): 281–90.

12. Chiotos K, Bassiri H, Behrens EM, et al. Multisystem Inflammatory Syndrome in Children During the Coronavirus 2019 Pandemic: A Case Series. J Pediatric Infect Dis Soc 2020; 9(3): 393–8.

13. Panupattanapong S, Brooks EB. New spectrum of COVID-19 manifestations in children: Kawasaki-like syndrome and hyperinflammatory response. Cleve Clin J Med 2020.

14. Ramcharan T, Nolan O, Lai CY, et al. Paediatric Inflammatory Multisystem Syndrome: Temporally Associated with SARS-CoV-2 (PIMS-TS): Cardiac Features, Management and Short-Term Outcomes at a UK Tertiary Paediatric Hospital. Pediatr Cardiol 2020.

15. Wang Y, Zhu F, Wang C, et al. Children Hospitalized With Severe COVID-19 in Wuhan. Pediatr Infect Dis J 2020; 39(7): e91–e4.

16. Richardson S, Hirsch JS, Narasimhan M, et al. Presenting Characteristics, Comorbidities, and Outcomes Among 5700 Patients Hospitalized With COVID-19 in the New York City Area. JAMA 2020.

17. Kim L, Whitaker, M., O’Halloran, A., et al. Hospitalization Rates and Characteristics of Children Aged <18 Years Hospitalized with Laboratory-Confirmed COVID-19 — COVID-NET, 14 States, March 1-July 25, 2020. MMWR Morb Mortal Wkly Rep 2020; 69.

18. Burke RM, Killerby ME, Newton S, et al. Symptom Profiles of a Convenience Sample of Patients with COVID-19 -United States, January-April 2020. MMWR Morb Mortal Wkly Rep 2020; 69(28): 904–8.

19. Gudbjartsson DF, Helgason A, Jonsson H, et al. Spread of SARS-CoV-2 in the Icelandic Population. N Engl J Med 2020; 382(24): 2302–15.

20. Davies NG, Klepac P, Liu Y, et al. Age-dependent effects in the transmission and control of COVID-19 epidemics. Nat Med 2020.

21. Posfay-Barbe KM, Wagner N, Gauthey M, et al. COVID-19 in Children and the Dynamics of Infection in Families. Pediatrics 2020.

22. Nashville COVID-19 Response: Safer at Home Order. Available at: https://www.asafenashville.org. Accessed July 7, 2020.

